# High Prevalence of Adolescents “At Risk for PCOS” and Links to Genetic Susceptibility to PCOS

**DOI:** 10.64898/2025.12.19.25342512

**Authors:** Varun Lingadal, Mali DiMeo, Joel N. Hirschhorn, Yee-Ming Chan, Jia Zhu

## Abstract

**Importance:** Diagnosis of polycystic ovary syndrome (PCOS) in adolescence is challenging because menstrual irregularity and hyperandrogenism are common in adolescents. Recent international guidelines highlighted an at risk for PCOS category based on either menstrual regularity or hyperandrogenism; however, its population prevalence and genetic correlates remain unknown.

**Objective:** To estimate the prevalence of PCOS and at risk for PCOS in adolescence and evaluate associations with genetic risk for PCOS.

**Design, Settings, and Participants:** Population-based analysis of 1,533 adolescents from the Avon Longitudinal Study of Parents and Children (ALSPAC) with sufficient reproductive data.

**Exposure:** Polygenic score (PGS) for PCOS derived from the largest genome-wide association study in European women.

**Main Outcomes and Measures:** Guideline-defined PCOS (presence of both irregular menses and hyperandrogenism) and at risk for PCOS (presence of one feature).

**Results:** PCOS prevalence was 3.2%, while 27.2% met criteria for being at risk for PCOS. A higher PCOS PGS was associated with hyperandrogenism (OR per SD increase in PGS: 1.22; 95% CI, 1.07-1.39; *P*=4×10^−3^) but not irregular menses.

**Conclusions and Relevance:** Over one-fourth of adolescents met criteria for being at risk for PCOS. Genetic risk for PCOS was associated with hyperandrogenism but not isolated menstrual irregularity, suggesting that androgen excess is a more specific early manifestation of inherited PCOS liability.

## Introduction

Polycystic ovary syndrome (PCOS) is a common endocrine disorder that often emerges during adolescence and carries lifelong reproductive and cardiometabolic consequences.^1^ Early identification is challenging because defining features of PCOS – menstrual irregularity and findings suggesting hyperandrogenism, such as acne – are common in adolescent girls. This creates diagnostic uncertainty and challenges in clinical counseling, monitoring, and early intervention.^2^

To address this challenge, 2025 international PCOS guidelines for adolescents emphasized persistent menstrual irregularity and androgen excess as the defining features of PCOS.^2^ Importantly, the guidelines highlight an “at risk for PCOS” category for individuals with only one feature, either irregular menses or hyperandrogenism. Emerging evidence suggests that adolescents in this group exhibit early metabolic alterations, including higher blood pressure and insulin resistance.^3,4^ However, the prevalence of this “at risk” category and its relationship to inherited biological factors that contribute to PCOS development remain unknown.

Using data from the Avon Longitudinal Study of Parents and Children (ALSPAC), a population-based birth cohort in the United Kingdom,^5^ we estimated the prevalence of the “PCOS” and “at risk for PCOS” classifications in adolescents and evaluated whether polygenic risk for PCOS is associated with these categories.

## Methods

Among 7,347 female children, 5,361 completed at least one reproductive questionnaire at age 8 years or later. After excluding individuals with premature ovarian insufficiency, pregnancy, contraception use during the assessment window, and incomplete data, 1,533 adolescents had sufficient data to assess both menstrual regularity and hyperandrogenism (Figure 1).

**Figure 1.**
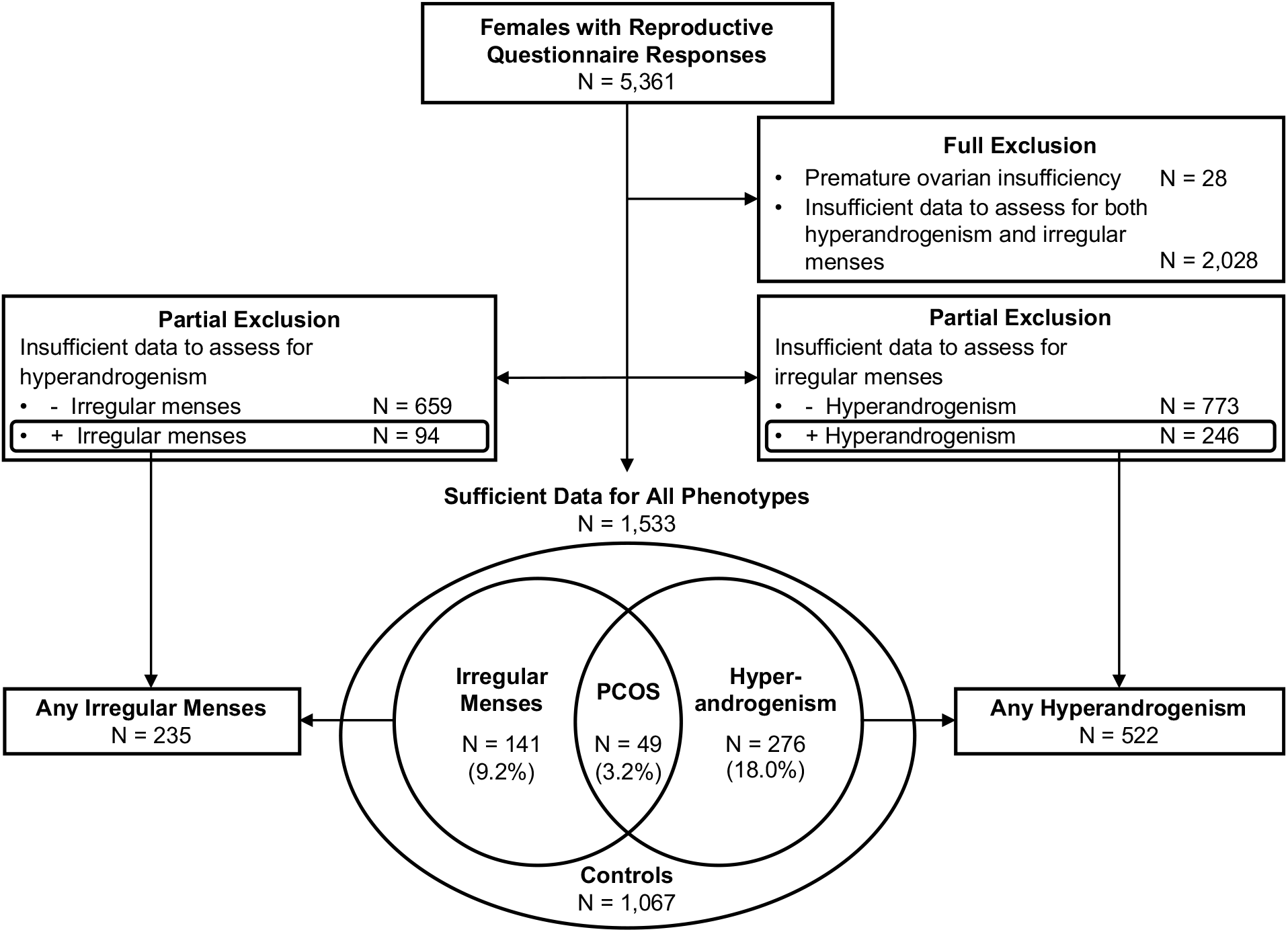
Flowchart of Classification of “PCOS” and “At Risk for PCOS” in Adolescent Females in ALSPAC. Calculations of the prevalence of the “PCOS” and “at risk for PCOS” categories were based on those with sufficient data for all phenotypes. Comparison of PCOS polygenic scores was done between the controls with sufficient data to exclude both irregular menses and hyperandrogenism and 1) the individuals meeting the criteria for PCOS, 2) the individuals meeting the hyperandrogenism criteria for “at risk for PCOS” (even if there were insufficient data to evaluate for irregular menses), and 3) the individuals meeting the irregular menses criteria for “at risk for PCOS” (even if there were insufficient data to evaluate for hyperandrogenism). Abbreviations: PCOS, polycystic ovary syndrome.

Menstrual irregularity was defined using age- and menarche-specific cycle-length criteria from the 2025 international guidelines. Hyperandrogenism was defined using biochemical measures (total testosterone and free-androgen index at age 15.5 years) and/or clinical hirsutism (modified Ferriman-Gallwey score ≥ 4 at age 19.5 years). “PCOS” classification required both features; “at risk for PCOS” required one feature without evidence of the other.^2^

Polygenic scores (PGS) for PCOS were calculated using PRS-CS and summary statistics from the largest GWAS meta-analysis for PCOS in European women.^6^ Logistic regression models, adjusted for the first 10 genetic principal components, tested associations between the PCOS PGS and the “PCOS” and “at risk for PCOS” classifications.

## Results

Among 1,533 female adolescents who had sufficient data to assess for all phenotypes, 49 (3.2%) met criteria for PCOS. An additional 417 (27.2%) met criteria for either irregular menses or hyperandrogenism and were classified as “at risk for PCOS,” with 276 (18.0%) having hyperandrogenism only and 141 (9.2%) having irregular menses only (Figure 1).

Among 1,371 genotyped adolescents of European ancestry, including those “at risk of PCOS” defined by any hyperandrogenism or irregular menses (Figure 1), a higher PCOS PGS was associated with increased odds of meeting the hyperandrogenism criterion (odds ratio [OR] per 1 SD increase in PGS 1.22; 95% CI, 1.07-1.39; *P*=4×10^−3^) but not the irregular menses criterion (OR 0.98; 95% CI, 0.83-1.15; *P*=0.8; Figure 2). The association between the PCOS PGS and the “PCOS” classification was positive but not significant (OR 1.28; 95% CI, 0.88-1.87, *P*=0.2), likely reflecting limited statistical power from low prevalence.

**Figure 2.**
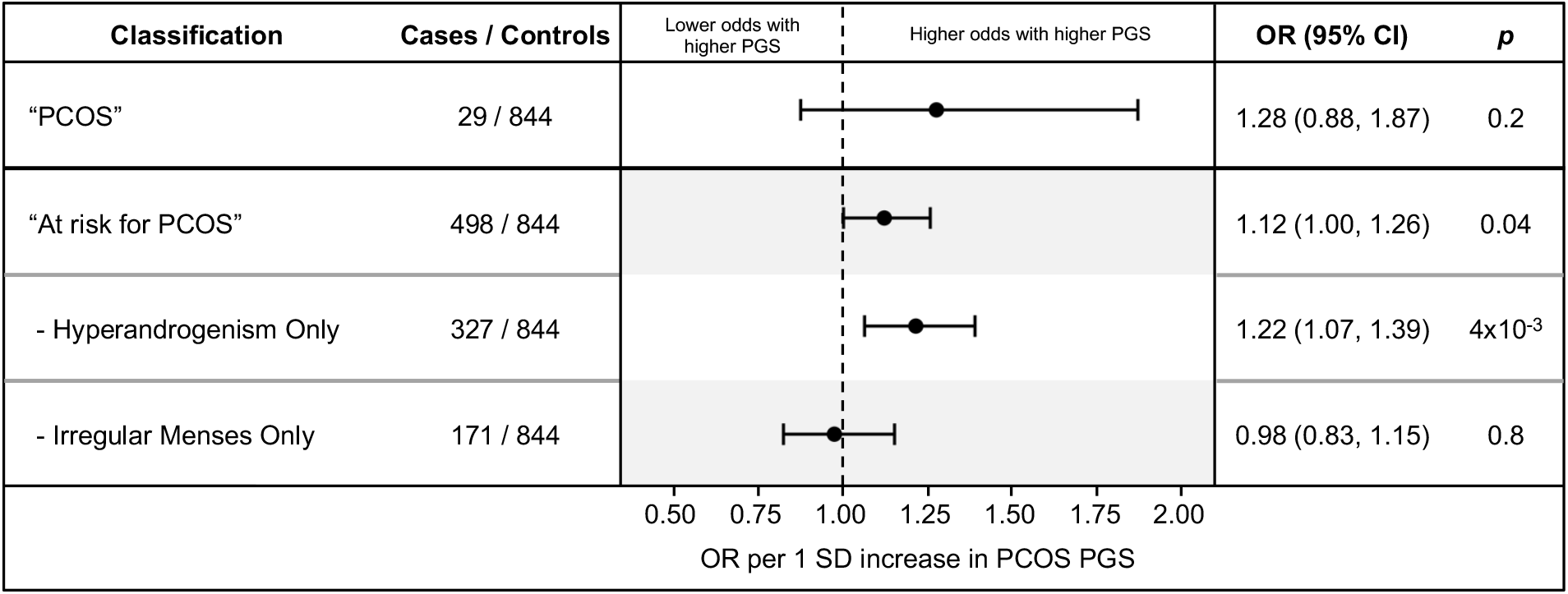
Relationships of PCOS Polygenic Score (PGS) with “PCOS” and “At Risk for PCOS.” Associations are shown for PCOS and for the “at risk for PCOS” classification overall and stratified by the defining criterion (hyperandrogenism or irregular menses). The “at risk for PCOS” category included any individual who met one criterion, including those with insufficient data to evaluate for the other criterion. Controls are individuals with sufficient data to exclude both irregular menses and hyperandrogenism. Abbreviations: PCOS, polycystic ovary syndrome; PGS; polygenic score; OR, odds ratio; CI, confidence interval.

## Discussion

In this population-based cohort of adolescents, PCOS prevalence was 3.2%, yet more than one-fourth met criteria for being “at risk for PCOS.” Higher polygenic risk for PCOS was associated with hyperandrogenism but not irregular menses, suggesting that hyperandrogenism represents a more specific early manifestation of inherited PCOS liability. In contrast, menstrual irregularity may either reflect a normal transitional stage during adolescence or aspects of PCOS pathophysiology that are not well captured by current GWAS-derived genetic scores. These findings support guideline recommendations for increased surveillance of adolescents with hyperandrogenism in particular and underscore the need for longitudinal studies to characterize progression from at-risk states to PCOS.

## Data Availability

Restrictions apply to the availability of all data generated during this study. Data from ALSPAC are available to all researchers, and data access is governed by the ALSPAC Executive Board of Directors (https://www.bristol.ac.uk/alspac/researchers/access/).

