## Supplemental Methods for "High Prevalence of Adolescents “At Risk for PCOS” and Links to Genetic Susceptibility to PCOS"

### Study Cohort

The Avon Longitudinal Study of Parents and Children (ALSPAC) is a population-based birth cohort study of 14,541 mother-child pairs.[1] Of these pregnancies, there were 13,988 children who were alive at 1 year of age and have been followed with longitudinal clinic- and questionnaire-based assessments.[1] At the time of analysis, 8,927 unrelated individuals of European ancestry had genotyping data available.

Individuals were considered lost to follow-up if they did not return the reproductive questionnaire. Data from returned questionnaires were assessed for completeness. For the menstrual regularity assessment, an incomplete classification was assigned if data on menstrual cycle length, age at menarche, and/or contraception use were not available. For the hyperandrogenism assessment, an incomplete classification was assigned if both data on labs (total testosterone and SHBG) or the hirsutism questionnaire were not available.

### "Irregular Menses" Classification:

Menstrual-cycle regularity was primarily assessed using questionnaire data on menstrual cycles, menarche status, and contraception use, which were completed approximately annually from 8 years to 17 years of age and at 19.5, 21, and 24 years (Supplemental Table 1).[2] Questionnaire responses were excluded if there was contraceptive use within 1 year prior to completion of the questionnaire. Responses were also excluded if the timing of pregnancy overlapped with the time period during which menses were classified as irregular. Participants with premature ovarian insufficiency, defined as reporting that periods stopped because of surgery, chemotherapy, radiation therapy, or menopause or for no apparent reason were also excluded.

"Irregular menses" was defined per the following criteria from the 2025 International evidence-based recommendations for polycystic ovary syndrome (PCOS) in adolescents: 1) onset of menarche after 15 years of age, 2) from 1 to <3 years post-menarche, mean or usual cycle length <21 days or >45 days, 3) from 3 or more years post-menarche, mean or usual cycle length <21 days or >35 days, and/or 4) after 1 year post-menarche, any one cycle >90 days. Age at menarche was determined based on reported age of first menstrual period.[2,3]

**"Hyperandrogenism" Classification**

Total testosterone and sex hormone-binding globulin (SHBG) were measured at approximately 15.5 years of age. The free-androgen index (FAI), a measure of androgen bioavailability, was calculated by dividing the total testosterone level (nmol/L) by the SHBG level (nmol/L) and multiplying by 100.[4] Biochemical hyperandrogenism was defined by a total testosterone and/or FAI greater than two standard deviations above the mean for females in the ALSPAC cohort. Because biochemical hyperandrogenism cannot be reliably assessed in individuals using combined oral contraceptive pills, we excluded participants who reported contraceptive use within the prior year.

Clinical hyperandrogenism was assessed using questionnaire data at 19.5 years. Modified Ferriman-Gallwey scores were calculated from participant responses on the presence and amount of unwanted/excess body hair in nine regions of the body (upper lip, chin, chest, upper back, lower back, upper abdomen, lower abdomen, legs, and arms).[5] Clinical hyperandrogenism was defined by a modified Ferriman-Gallwey score $\geq 4$.[6] Participants who were pregnant at the time of assessment were excluded because pregnancy can influence hirsutism.[7]

**PCOS Genetic Risk Score Calculation and Analysis**

Polygenic scores (PGS) for PCOS were calculated using PRS-CS and summary statistics from the largest GWAS meta-analysis for PCOS in European women.[8] This study included data from 20,818 cases and 523,695 controls, and 93% of the cohort was of European ancestry. PRS-CS software was used to generate a PGS for PCOS using a Bayesian approach that adjusts for linkage disequilibrium by using a reference panel and weights the effect size of each variant by the strength of its association in GWAS, which allows inclusion of all variants.[12] Our PCOS PGS was calculated using 1,119,009 genetic variants and was scaled to a mean of 0 and an SD of 1 to facilitate interpretation.

For comparison of PCOS PGS between individuals "at risk for PCOS" to controls, the analysis was performed in two ways. For the primary analysis, individuals with sufficient data to evaluate for hyperandrogenism or menstrual irregularity were included in analysis even if they had insufficient data to evaluate the other PCOS feature. This approach risks including individuals meeting full PCOS criteria in one of these "at risk for PCOS" categories. To assess for possible bias from this potential misclassification, we conducted a sensitivity analysis restricted to adolescents with sufficient data to assess for both hyperandrogenism and irregular menses, and we found effect sizes comparable to those found in the primary analysis.

Supplemental Table 1. ALSPAC G1 Patient Source Variables

| **Variable Type** | | Menstrual Cycle Length | Contraception Use / Type | Menarche & Period Status | Pregnancy / Parent Status |
|---|---|---|---|---|---|
| **Age Group Questionnaires** | | | | | |
| Growing and Changing | 8y 1m | pub117 | pub127 | clon070, pub710, pub810, pub910 | NA |
| | 9y 7m | pub217 | pub227 | | NA |
| | 10y 8m | pub317 | pub327 | | NA |
| | 11y 8m | pub417 | pub427 | | NA |
| | 13y 1m | pub517 | pub527 | | NA |
| | 14y 7m | pub617 | pub627 | | NA |
| | 15y 6m | pub717 | pub727 | | NA |
| | 16y 0m | pub817 | pub827 | | NA |
| | 17y 0m | pub917 | pub927 | | NA |
| Teen Focus 4: Focus at 17 | 17y 0m | NA | FJMS010, FJMS011, FJMS012, FJMS013, FJMS014 | FJMS020 | NA |
| You and Your Body Aged 19+ | 19y 6m | ccxf3000 | ccxf2000, ccxf2001, ccxf2002, ccxf2003, ccxf2004, ccxf3004 | NA | ccxf4000, ccxf4001, ccxf4002, ccxf5000 |
| Your Life Now (21+) | 21y | NA | NA | NA | NA |
| Focus @ 24 | 24y | FKFH1020 | FKFH1040 | NA | FKFH1071 |
| Life @ 24+ | 24y | NA | NA | NA | YPD2040 |
